# Short Adaptation of the Smell Test of the University of Pennsylvania (UPSIT) For the Venezuelan Population

**DOI:** 10.1101/2020.08.25.20182063

**Authors:** Pieruzzini Rosalinda, Rodríguez Wilneg, Velázquez Carlos

## Abstract

The objective of this study was to design a short smell test adapted to the Venezuelan population using the UPSIT test as a reference.Methodology: Preliminary surveys were carried out in 2 groups of patients (1010 patients for each group) until obtaining the most frequently recognized odors and preferred values of that population. Normosmia, hyposmia and anosmia were normalized. The results of the new Venezuelan test were compared with the UPSIT test in 169 patients. Results: Sensitivity 73%, Specificity 100% and Positive predictive value of 100 with a diagnostic accuracy of 85,21%. Conclusions: short venezuelan smell test is safe and with reliable results in the diagnosis of olfactory alterations.

## INTRODUCTION

Smell is phylogenetically considered one of the oldest senses, being less specialized in man in relation to other lower species. Smells impact our emotional life and through the sense of smell we can associate and evoke feelings. Smell are the sense of attraction, good spirits, memory, relaxation and sensuality; is a vital gift and plays a very important role in the quality of life of the human being, in eating habits, in nutrition, in interpersonal relationships; in addition, it informs us of situations of danger to life.

Olfactory dysfunction is a very common condition with a prevalence between 4 and 25%. Men are more likely to suffer these alterations compared to women, especially if they are exposed to smoking or work in industrial areas.^(1)^ In Venezuela these alterations are a little bit diagnosed because they are not done intentionally, as well as because of the scarcity training of medical personnel on the subject, therefore it is not easy to specify the real prevalence of smell disorders in our country since it is not considered a mandatory report disease, there is no real casuistry registered and in turn it does not constitute a sole reason for query. The detection and diagnosis of olfactory disorders is based on a detailed clinical history at the general and otorhinolaryngological level, as well as on the questioning and physical examination, followed by the performance of a smell test ^(2)^. Doctors and researchers have developed different types of tests that allow assessing olfactory function as well as determining the nature and degree of smell disorders in people. Among these are olfactometries. Olfactometry is the measurement of olfaction using the set of tests that study olfactory function. These are intended to find a threshold of olfaction (liminal olfactometry) to establish quantitative alterations and study disturbances above the threshold (supraliminal olfactometry) and thus reveal qualitative alterations’ ^(3)^

The tests that are based on the appraisals of the explored individual are called subjective, they have the disadvantage of being influenced by psychic, gustatory and trigeminal factors; which can influence the results, the objectives are based on the detection of changes in the central nervous system caused by olfactory stimuli, useful in patients who cannot collaborate or in simulators, they record the response of the individual being explored without participating with it. your interpretation of the feeling. ^(3)^ Unfortunately, most of the validated odor identification tests are not cross-cultural, and those that are currently commercially available have been developed in the study of populations from the United States of America, Europe, and Asia. ^(4)^

The UPSIT (University of Pennsylvania Smell Identification Test): It is the most widely used olfactory test ^(5)^. This test consists of four “scratch and smell” booklets that require no training, can be self-administered, or can be administered by a caregiver, nurse practitioner, or physician. The UPSIT has been used in clinical and research settings and has proven valuable in diagnosing loss of smell due to neurological disease, as well as for primary ENT conditions, such as chronic sinusitis, among others. It has also been used to aid in the differential diagnosis of parkinsonism ^(6)^ and dementia ^(7)^ In Brazil, a Brazilian-Portuguese version of UPSIT has been developed, called UPSIT-Br2, in which the elements and response options that were considered inappropriate for Brazilians (because they did not recognize some substances) were replaced by elements and options that are considered more familiar to them, obtaining satisfactory results, to normalize this test in Brazil ^(8)^. A subject with normosmia is considered when the odor detection is at 38-40. In view of this antecedent, it was decided to carry out a research protocol aimed was, perform a short smell test that will be adapted to the needs of the Venezuelan population, taking the UPSIT test as a reference point and in order to avoid generating false positives in the results.

## Methods

Population and sample: The sample was constituted for 2020 Venezuelan subjects over 18 years of age, without disorders of the otorhinolaryngological or neurological sphere, randomly chosen for the phases of the test development. The validation phase was carried out with 118 individuals who attended the ENT consultation.

### Procedure

A survey was designed as a data collection instrument, emphasizing age, sex, direction, knowledge and preference of odoriferous substances in order to characterize the Venezuelan population. The application of this instrument was carried out in 5 phases, which are described below.

1st Phase: Survey application in 1010 volunteer subjects. In this first phase, the subject is informed which are the classic substances of the UPSIT test to determine if he recognizes them or has heard them named and in a second part of the survey, the subject is indicated some typical smells of the culture of the Venezuelan to compare their recognition capacity between both types of odoriferous substances. Those substances unknown to the patients and those that caused confusion were identified, marking with an “X” (X) or a “check mark” those known. The third part corresponds to suggesting an odoriferous substance and the fourth part indicates a favorite odoriferous substance of the respondent.

2nd Phase: Preparation of a provisional smell test with results of the survey. A total of 10 odorants were presented, which were placed in plastic bottles with a capacity of 1210 ml, with an opening of 4.5 cm in diameter and 6 cm deep, without identification. The smells presented were the following: coffee, chocolate, coconut, baby cologne, liquid disinfectant, baby powder, washing powder, cinnamon, acetone and rum. This test was prepared at home, acquiring the substances prepared as essences in commercial houses: 50 ml of coconut essence, baby cologne, liquid disinfectant, acetone and rum. 20 grams of coffee powder, cinnamon, chocolate, ACE, talc. Each odorant was presented with an interval of 45 seconds, which is the minimum time for adaptation of the olfactory receptors. The odors were presented in bottles without identification and with the eyes occluded and presenting the substance 1 cm from the nostrils, which were evaluated separately, occluding one nostril and then the other. For the identification of the odorants in the test, a leaflet was presented showing the exposed odoriferous substance plus 3 distractors. The participants could choose one of the 4 options presented to identify each odorant. Of the 10 smells tested, the individual had to recognize the majority of odorants to validate the test in the population studied. It is accepted that the recognition of between 8 and 10 smells, the subject was normosmic.

3rd Phase: Application of the provisional smell test, in 1010 subjects, volunteers, chosen at random, under the same conditions as the first ones, in order to eliminate any possibility of bias of the researcher and the population exposed to the modified test, since by knowing the substances and the olfactory memory capacity, they could easily identify the smells chosen by them. Upon completion of the test, the examiner had to be sure that the identification of the 10 odorants was correct.

4th Phase: The smell test adapted to the Venezuelan population was prepared, which has a card to empty personal data, an instructions card and an answer card, in addition to 2 notebooks with the 10 odorants selected based on the results obtained in the survey and approved after the application of the provisional test. Each of the notebooks has 5 pages and each page you will find the chosen odorant and the name of 3 distracting substances.In this phase was made the normalization of the test.

5^th^ Phase: Validation of the Venezuelan test comparing it with the UPSIT in 169 patients who consulted the otolaryngology triage.

### Bioethical approval

All participating subjects were requested authorization to participate in the study through informed consent and the protocol was evaluated and approved by the Bioethics Commission of the Hospital Militar Universitario “Dr. Carlos Arvelo ”. All data and information collected were used only for the purpose of research.

Analysis of data: Once the data were collected, they were systematized in a master table in Microsoft® Excel and then processed, presented and analyzed using descriptive and inferential statistical techniques, in tables of frequency and association distributions, according to the specific objectives proposed. For the age variable, once its tendency to normality had been demonstrated, the mean ± standard error, median, minimum value, maximum value and coefficient of variation were calculated, comparing it according to sex using the hypothesis test for the difference between means (t student). The variable quantity of recognized odors, being discrete, was calculated median, interquartile range, minimum and maximum value, compared according to age groups using the Kruskall Wallys and Mann Whitney W tests to compare medians. The quantity of recognized odors according to age groups and sex was associated (related) by means of the non-parametric Chi-square analysis for independence between variables. Everything was carried out using the SPSS statistical processor for Windows version 20, adopting P values lower than 0.05 (P <0.05) as the level of statistical significance. The sensitivity and specificity were calculated using the Wilson points method with a 95% confidence interval, and a corrected Kappa index was applied.

## Results

Of the 1010 subjects that made up the sample, an average age of 41.85 ± 0.56 years was recorded, with a median of 37 years, a minimum age of 18 years, a maximum age of 94 years and a coefficient of variation of 43% (series moderately heterogeneous among their data). Those subjects aged 18 and 27 years (27.92% = 282 cases) predominated, followed by those aged 28 and 37 years (22.48% = 227 cases). The female sex represented 60% of the sample under study (606 cases), while the male represented 40% (404 cases). There was no statistically significant difference between the averages of age according to sex (t = 0.18; P = 0.8576> 0.05). (Table no1)

**Table No. 1.** Characterization according to age and sex of the sample of the Venezuelan population interviewed, for the proposal of the smell evaluation test adapted to the Venezuelan population.

| Age | f | % |
| --- | --- | --- |
| 18 – 27 | 282 | 27,92 |
| 28 – 37 | 227 | 22,48 |
| 38 – 47 | 110 | 10,89 |
| 48 – 57 | 182 | 18,02 |
| 58 – 67 | 130 | 12,87 |
| 68 – 77 | 38 | 3,76 |
| $\geq 78$ | 41 | 4,06 |
| Average | $\bar{X} \pm Es$ | |
| Total sample | 41,85 años ± 0,56 | |
| Female | 41,95 años ± 1,44 | |
| Male | 41,73 años +/- 1,71 |  |
| **Sex** | **f** | **%** |
| Female | 606 | 60 |
| Male | 404 | 40 |
| **Totally** | **1010** | **100** |

All the subjects that made up the study sample (1010 cases), agreed in the identification of the smell of coffee, chocolate, Lavansan disinfectant, cinnamon, ACE detergent, coconut, talcum powder, rum, small colony, and acetone. In addition to the unanimous identification of the aforementioned odors, another recognized odorant was orange (90.69% = 916 cases) and nail painting (75.34% = 761 cases). The unrecognized odors were Lilac, Turpentine, Moss and Tire, which belong to the UPSIT test. (table no2)

**Table nº2.** Recognized odour on the studied population

| Recognized | Yes |  | No |  |
| --- | --- | --- | --- | --- |
| Odour | F | % | f | % |
| Coffee | 1010 | 100 | 0 | 0 |
| Chocolate | 1010 | 100 | 0 | 0 |
| Disinfectant | 1010 | 100 | 0 | 0 |
| Cinnamon | 1010 | 100 | 0 | 0 |
| Washing powder | 1010 | 100 | 0 | 0 |
| Coconut | 1010 | 100 | 0 | 0 |
| Baby powder | 1010 | 100 | 0 | 0 |
| Rum | 1010 | 100 | 0 | 0 |
| Baby Cologne | 1010 | 100 | 0 | 0 |
| Acetone | 1010 | 100 | 0 | 0 |
| Orange | 916 | 90,69 | 94 | 9,30 |
| Nail Polish | 761 | 75,34 | 249 | 24,65 |
| Alcohol | 734 | 72,67 | 276 | 27,32 |
| Grama | 669 | 66,23 | 341 | 33,76 |
| Watermelon | 588 | 58,22 | 422 | 41,78 |
| Pinesol | 528 | 52,27 | 482 | 47,7 |
| Tinner | 367 | 36,34 | 643 | 63,66 |
| Passion fruit | 358 | 35,44 | 652 | 64,55 |
| Cigarette | 124 | 12,27 | 886 | 87,72 |
| Peanut | 108 | 10,69 | 902 | 89,30 |
| Lilac | 0 | 0 | 1010 | 100 |
| Turpentina | 0 | 0 | 1010 | 100 |
| Moss | 0 | 0 | 1010 | 100 |
| Tire | 0 | 0 | 1010 | 100 |

Of the 1010 subjects included in the sample, a median of 17 recognized odors was recorded, with an interquartile range of 2 odors, a minimum record of 13 recognized odors, and a maximum record of 20 recognized odors. A statistically significant difference was found between the medians of recognized odors according to the age groups (KW = 27.25; P = 0.0001 <0.05), the lowest median being that registered by those subjects aged 68 and 77 years. According to sex, no statistically significant difference was found between the medians (W = 124 989.0; P = 0.5624> 0.05). Those subjects who recognized between 17 and 20 odors from the test designed by the researchers predominated, representing 62.38% of the sample studied (630 cases). According to the age groups, it was the subjects aged 18 and 27 who recognized the most odors (160 cases), followed by those aged 48 and 57 (136 cases). Of the people who recognized the least amount of odors, those aged 18 and 27 were the most frequent age group (122 cases). A statistically significant association was found between the amount of recognized odors and the age groups (X2 = 28.47; 6 gl; P = 0.0001 <0.05). According to sex, it can be said that both sexes registered a similar proportion in terms of the amount of recognized odors: female (374/606) and male (256/404). No statistically significant association was found between the amount of recognized odors and sex (X2 = 0.22; 1 gl; P = 0.6426> 0.05). (table 3)

**Table 3.** Recognized Odour, age and sex

| RECOGNIZED ODOUR | 13 – 16 |  | 17 – 20 |  | Totally |  | Xd - RI |
| --- | --- | --- | --- | --- | --- | --- | --- |
| Age | f | % | f | % | f | % |  |
| 18 – 27 | 122 | 12,08 | 160 | 15,84 | 282 | 27,92 | 17 – 2 |
| 28 – 37 | 101 | 10 | 126 | 12,48 | 227 | 22,48 | 17 – 2 |
| 38 – 47 | 30 | 2,97 | 80 | 7,92 | 110 | 10,89 | 17 – 2 |
| 48 – 57 | 46 | 4,55 | 136 | 13,47 | 182 | 18,02 | 17 – 2 |
| 58 – 67 | 45 | 4,46 | 85 | 8,42 | 130 | 12,87 | 17 – 3 |
| 68 – 77 | 19 | 1,88 | 19 | 1,88 | 38 | 3,76 | 16,5 – 2 |
| ≥78 | 17 | 1,68 | 24 | 2,38 | 41 | 4,06 | 17 – 2 |
| Sex | f | % | f | % | f | % |  |
| Female | 232 | 22,97 | 374 | 37,03 | 606 | 60 | 17 – 2 |
| Male | 148 | 14,65 | 256 | 25,35 | 404 | 40 | 17 – 2 |
| **Totally** | **380** | **37,62** | **630** | **62,38** | **1010** | **100** |  |

The first 10 odoriferous substances were chosen from among the suggested and preferred substances and the provisional test was constructed. After applying it, 100% of the substances were recognized in the subjects that made up the pilot sample (1010 cases).(Table 4)

**Table.4.** Provisional Test with definitive odorants

| Recognized | Yes |  | No |  |
| --- | --- | --- | --- | --- |
| Olores | f | % | f | % |
| Washing Powder | 1010 | 100 | 0 | 0 |
| Cinnamon | 1010 | 100 | 0 | 0 |
| Coffee | 1010 | 100 | 0 | 0 |
| Chocolate | 1010 | 100 | 0 | 0 |
| Baby powder | 1010 | 100 | 0 | 0 |
| Rum | 1010 | 100 | 0 | 0 |
| Disinfectant | 1010 | 100 | 0 | 0 |
| Coconut | 1010 | 100 | 0 | 0 |
| Acetone | 1010 | 100 | 0 | 0 |
| Baby cologne | 1010 | 100 | 0 | 0 |

### Normalization of the Venezuelan test

The number of correct answers presented a Gaussian distribution, according to the Kolmogorov-Smirnov normality test (median: 10, standard deviation: 1.49). From this distribution, the normality / hyposmia / anosmia values were defined using the standard deviation . Of the total number of patients (169), 102 patients obtained a score between 8 and 10 (60,35%), 49 patients obtained a score between 6 and 7 (28,99%),11 patients (6,50%) obtained a score between 4 and 5, 3 patients(1,77%) obtained a score between 2 and 3 and 4 patients (2,36%) between 0 and 1.

Normosmia: 8-10 Mild Hyposmia: 6-7 Moderate Hyposmia: 4-5 Severe Hyposmia: 2-3 Anosmia: 0-1

**Table 5.** Sensitivity and specificity of Venezuelan Test.Predictive Value

|  |  | UPSIT<br>Positive | UPSIT<br>Negative | Totally |
| --- | --- | --- | --- | --- |
| Venezuelan | Positive | 67 | 0 | 67 |
| Test | Negative | 25 | 77 | 102 |
|  | Totally | 92 | 77 | 169 |

The Venezuelan Test sensitivity is 72,83% and specificity 100%. Positive Predictive Value 100%; Negative Predictive Value 75,49%,diagnostic accuracy 85,21%, Kappa Cohen’s 0,7095.(IC 95%)

## Discussion

The growing interest regarding the physiological and pathological processes that affect the sense of smell and its widely demonstrated correlation with neurodegenerative diseases, have led to the development of various techniques to objectify its deficit in order to use it in the early detection of these diseases ^(9)^. In our study, the identification of ten odorants in a healthy Venezuelan population is evaluated using a test adapted to the population, based on the UPSIT test, which, to date, was based on normal values of olfactory sensitivity in healthy patients from the American population., European or Asian.

The population analyzed in our study shows a higher performance in the recognition of coffee, chocolate, coconut, washing powder,cinnamon,baby powder,rum,disinfectant,acetone and baby cologne. The performance of odor recognition decreases when options are presented to describe or remember such as tire, turpentine, lilac and moss, which is due to a cultural factor, since they are unfamiliar aromas in our environment. Previous studies with the use of this and other techniques have determined that olfaction levels decrease with age in both sexes and that men under physiological conditions smell less than women. ^(10)^ Our study visualizes the fact that that both genders identify substances equally, the variations between the groups did not show statistically significant differences, however, it is observed that in the age groups between 18 and 27 years of age there is a greater recognition of odoriferous substances than in the others groups. With the results obtained from the interview, 1010 volunteer subjects were taken, taken randomly, under the same conditions as the first ones, in order to eliminate any possibility of bias of the researcher and of the population exposed to the modified test, since knowing the substances and thanks to the olfactory memory capacity they could easily identify the smells chosen by them, we proceeded to the application of the provisional test of the evaluation of smell, it was obtained that the recognition of the selected odoriferous substances was 1010 (100%) in the subjects that made up the pilot sample. After the design of the smell evaluation test adapted to the Venezuelan population based on the smell identification test of the University of Pennsylvania, it was applied, obtaining the recognition of 100% of the odoriferous substances selected by the sample. Taking as reference the study was leaded by Hudson et al.^10^ in the study was leaded in Chile, established the normalization of the values obtained when applying the smell test in 99 healthy subjects. The number of correct answers presented a Gaussian distribution, according to the Kolmogorov-Smirnov normality test. From this distribution, the normality / hyposmia / anosmia values were defined using the standard deviation. Of the total number of patients, 68 patients obtained a score between 8 and 12 (69%), 29 patients obtained a score between 6 and 7 (29%) and 2 patients obtained a score less than or equal to 5 (2%). In our study, was similar behavior in the distribution of the data.

Validation of the Venezuelan test was performed with the UPSIT test and a sensitivity of 72.83% and specificity of 100% were obtained with a positive predictive value of 100% and a diagnostic precision of 85.21%, which allows us to affirm that its application in the the Venezuelan population has considerable evidence to make accurate diagnoses of olfactory disorders.

The application of test using the suggested method is easy, fast and eliminates the risk of contamination with the aroma of the fingers or hands of the researcher and the patient. This allows the measurement of smell at a supra-threshold level, which makes this test very favorable to be used as a rapid measurement or screening test.

## Conclusions

The smell test adapted to the Venezuelan population based on the odor identification test of the University of Pennsylvania is reliable and easy to apply in the office, being easy to use for both the examiner and the examinee, and the most important thing is that It can be used to evaluate the sense of smell in the Venezuelan population, since the odoriferous substances tested have a high degree of identification in this environment. The selection of odors was based on national cultural indicators, obtained through an interview by means of an evaluation instrument, it may be modified according to the clinical evaluation that arises from its use. This test is able to reliably differentiate patients with olfactory disorders from healthy subjects.

## Data Availability

All data is avalaible

## Notes

### Competing Interest Statement

The authors have declared no competing interest.

### Funding Statement

there was no financing

### Author Declarations

This investigation has been approved for the Bioethical Committee of Hospital Militar Universitario "Dr.Carlos Arvelo" Caracas-Venezuela.

